# Severity of maternal SARS-CoV-2 infection and perinatal outcomes during the Omicron variant dominant period: UK Obstetric Surveillance System national cohort study

**DOI:** 10.1101/2022.03.07.22271699

**Authors:** Hilde Marie Engjom, Rema Ramakrishnan, Nicola Vousden, Kathryn Bunch, Edward Morris, Nigel Simpson, Chris Gale, Pat O’Brien, Maria Quigley, Peter Brocklehurst, Jennifer J Kurinczuk, Marian Knight

## Abstract

**Objectives:** To describe the severity of maternal infection when the Omicron SARS-CoV-2 variant was dominant (15/12/21-14/01/22) and compare outcomes among groups with different vaccination status.

**Design:** Prospective cohort study

**Setting:** UK consultant-led maternity units

**Participants:** Pregnant women hospitalised with a positive SARS-CoV-2 PCR test up to 7 days prior to admission and/or during admission up to 2 days after giving birth.

**Main outcome measures:** Symptomatic or asymptomatic infection. Vaccination status. Severity of maternal infection (moderate or severe infection according to modified WHO criteria). Mode of birth and perinatal outcomes.

**Results:** Out of 1561 women admitted to hospital with SARS-CoV-2 infection, 449 (28.8%) were symptomatic. Among symptomatic women admitted, 86 (19.2%) had moderate to severe infection; 51 (11.4%) had pneumonia on imaging, 62 (14.3%) received respiratory support, and 19 (4.2%) were admitted to the intensive care unit (ICU). Three women died (0.7%). Vaccination status was known for 383 symptomatic women (85.3%) women; 249 (65.0%) were unvaccinated, 45 (11.7%) had received one vaccine dose, 76 (19.8%) had received two doses and 13 (3.4%) had received three doses. 59/249 (23.7%) unvaccinated women had moderate to severe infection, compared to 10/45 (22.2%) who had one dose, 9/76 (11.8%) who had two doses and 0/13 (0%) who had three doses. Among the 19 symptomatic women admitted to ICU, 14 (73.7%) were unvaccinated, 3 (15.8%) had received one dose, 1 (5.3%) had received two doses, 0 (0%) had received 3 doses and 1 (5.3%) had unknown vaccination status.

**Conclusion:** The risk of severe respiratory disease amongst unvaccinated pregnant women admitted with symptomatic SARS-CoV-2 infection during the Omicron dominance period was comparable to that observed during the period the wildtype variant was dominant. Most women with severe disease were unvaccinated. Vaccine coverage among pregnant women admitted with SARS-CoV-2 was low compared to the overall pregnancy population and very low compared to the general population. Ongoing action to prioritise and advocate for vaccine uptake in pregnancy is essential.

**SUMMARY BOX**
**What is already known on this topic**

- In non-pregnant adults, growing evidence indicates a lower risk of severe respiratory disease with the Omicron SARS-CoV-2 Variant of Concern (VOC).
- Pregnant women admitted during the periods in which the Alpha and Delta VOC were dominant were at increased risk of moderate to severe SARS-CoV-2 infection compared to the period when the original wildtype infection was dominant.
- Most women admitted to hospital with symptomatic SARS-CoV-2 infection have been unvaccinated.

**What this study adds**

- One in four women who had received no vaccine or a single dose had moderate to severe infection, compared with one in eight women who had received two doses and no women who had received three doses
- The proportional rate of moderate to severe infection in unvaccinated pregnant women during the Omicron dominance period is similar to the rate observed during the wildtype dominance period
- One in eight symptomatic admitted pregnant women needed respiratory support during the period when Omicron was dominant

## INTRODUCTION

In 2020 the World Health Organization’s (WHO) living systematic review concluded that SARS-CoV-2 infection during pregnancy was associated with an increased risk of admission to intensive care (ICU) for the mother, increased risk of preterm birth and admission for neonatal care for the infant.^1^ Included studies initially contained data predominately from the USA and China, with few active population-based surveillance studies. The last update was published in April 2021 and consequently included studies comprise mainly the Variants of Concern (VOC) prior to Delta.

In the UK, a new SARS-CoV-2 VOC (B.1.1.529, Omicron) was initially reported in November 2021 and dominated by mid-December 2021.^2^ With the Alpha (B.1.1.7) and Delta (B.1.617) VOC, severe maternal infection was more frequent compared to the wildtype period, and perinatal outcomes were worse^3-5^. The majority of severe maternal and perinatal outcomes occurred among unvaccinated women during periods with alpha and delta as dominant variants.^6 7 8^ Initial studies of Omicron infection in non-pregnant populations indicate a lower risk of severe pulmonary disease with this variant compared to the previous delta VOC.^9-11^

To date we have not identified any peer-reviewed published studies exploring the impact of infection with the Omicron SARS-CoV-2 variant on pregnant women and perinatal outcomes. There is an urgent need for robust national data to inform women who are pregnant or plan a pregnancy, as well as health professionals providing care for pregnant women, and policy makers. The primary aim of this study was therefore to describe the characteristics of pregnant women admitted to hospital with SARS-CoV-2 infection including vaccination status, severity of infection, pharmacologic management, pregnancy, and perinatal outcomes, in the first period when the Omicron VOC was dominant in the UK.

## METHODS

### Design, data sources and study period

A national, prospective observational cohort study was conducted using the UK Obstetric Surveillance System (UKOSS).^12^ This system entails active surveillance with reporting from all 194 hospitals in the UK with a consultant-led maternity unit, and includes well established routines to secure complete reporting.^13^ Information on women who died, or who had stillbirths or neonatal deaths, was cross-checked with data from the organisation responsible for maternal and perinatal death surveillance in the UK (MBRRACE-UK).^14^ As individual-level SARS-CoV-2 variant data were not recorded in medical records, the data collection time period was restricted to the period in which the Omicron SARS-CoV-2 variant was the dominant circulating strain in the UK. The cut-off at December 15 was chosen since the variant then represented 50% or more of sequenced new cases from Public Health England.^11^

### Study population and study groups

Women were included if they were admitted to hospital during pregnancy and had a positive SARS-CoV-2 PCR test at the time of admission. Hospital admission was defined as an overnight or longer hospital admission for any cause, or admission of any duration to give birth. Women not meeting this case definition were excluded (Figure 1). The included women were categorised in two mutually exclusive groups based on covid-19 symptoms. Symptomatic group: women who were admitted due to covid-19 or who were reported to be symptomatic or who received respiratory support of any kind.

**Figure 1.**
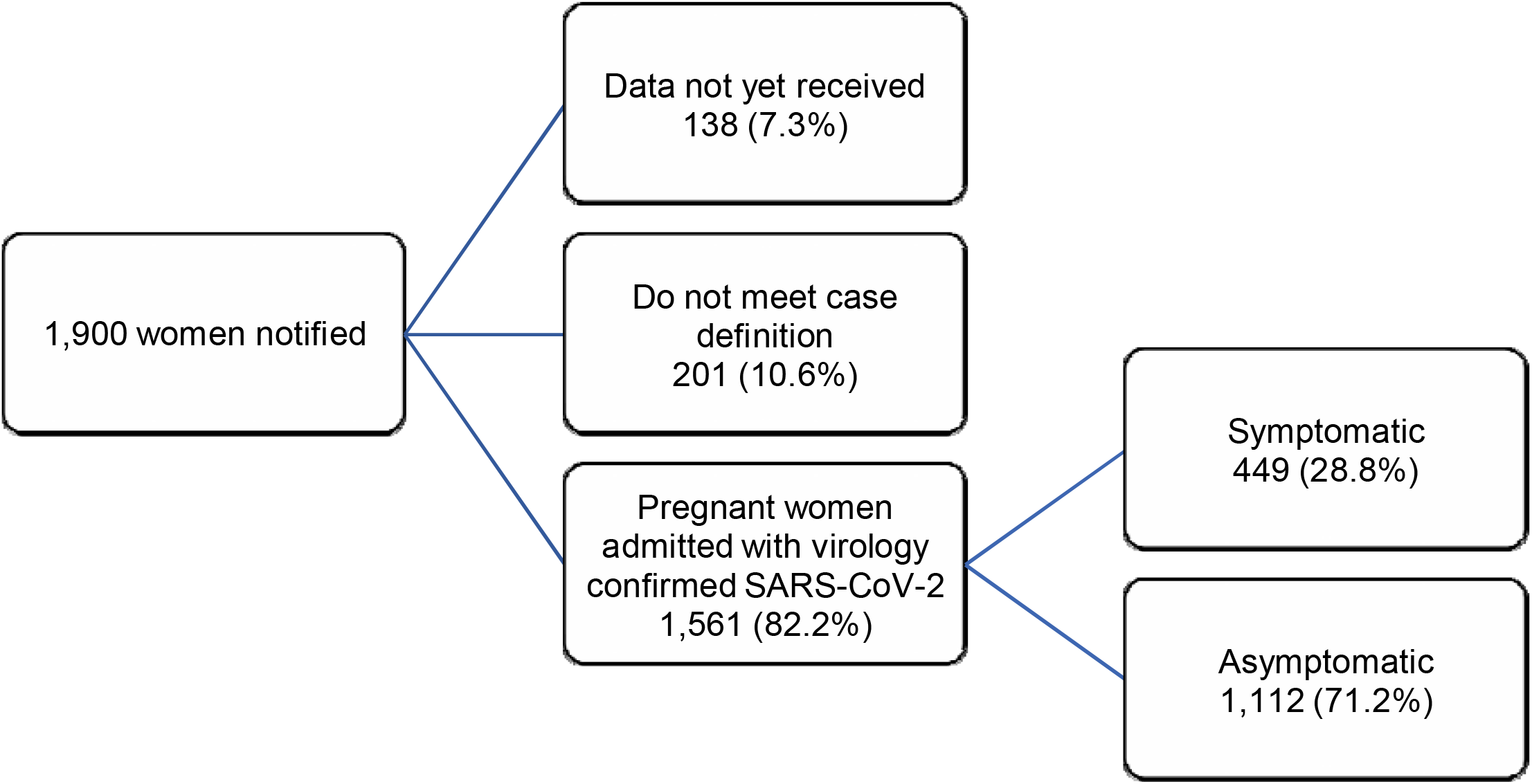
Inclusion flow chart: pregnant women admitted to hospital with SARS-CoV-2 infection, by admission group, UK, 15^th^ December 2021 to 14^th^ January 2022

Asymptomatic group: women admitted for labour, obstetric care or other reasons and who were not reported to have SARS-CoV-2-related symptoms and who did not receive respiratory support, or who were reported to be asymptomatic if the reason for admission was not known.

Vaccination status was categorised as follows; unvaccinated, one dose, two doses and three doses, or vaccination status unknown.

### Measures

A composite measure indicating moderate to severe SARS-CoV-2 infection was based on the WHO criteria of covid-19 disease severity.^15^ Women were classified as having moderate to severe respiratory disease if one or more of the following was reported: oxygen saturation <95% on admission, need for respiratory support, evidence of pneumonia on imaging, admission to ICU, or maternal death. Respiratory support was recorded as the maximum level of support in one of the following categories: oxygen therapy (supplementation on nasal prongs or non-rebreathe mask <15 l/min), high-flow nasal cannula ≥15 l/min or continuous positive airway pressure (CPAP), mechanical ventilation (MV) or extracorporeal membrane oxygenation (ECMO). Mode of birth was categorised as follows; caesarean section prior to or in labour, assisted vaginal birth, unassisted vaginal birth. Gestational age was categorised by completed weeks as <22 weeks, 22 to 27 weeks, 28 to 33 weeks, 34 to 36 weeks and ≥37 weeks for gestational age at admission and gestational age at childbirth. The following perinatal outcomes were included: total births, live births, stillbirths, neonatal unit (NNU) admission, early neonatal death.

The following sociodemographic and medical risk factors were included: maternal age, body mass index (BMI) in kg/m^2^, occupation (woman or partner in paid work vs neither in paid work), minority ethnic background (Asian, Black, Chinese, other or mixed ethnic minorities vs White), smoking (current smoker vs non-smoker), medical conditions prior to or during pregnancy (asthma, hypertension, cardiac disease, and diabetes prior to or in pregnancy), parity (nulliparous vs multiparous), plurality (singleton vs multiple).

Descriptions of pharmacological therapies were based on national guidance on pharmacological therapy issued on 01/07/2020 using the current edition at the time of admission^16^.

### Study registration

The study was registered with ISRCTN, number 40092247 and the protocol is available at https://www.npeu.ox.ac.uk/ukoss/current-surveillance/covid-19-in-pregnancy.

### Ethics and consent

This study was approved by the HRA NRES Committee East Midlands – Nottingham 1 (Ref. Number: 12/EM/0365).

### Patient and Public Involvement

Patients and public were part of the UKOSS steering committee and involved in study oversight but not in the design, reporting, conduct or dissemination of this study.

### Statistical methods and analysis

Continuous variables were summarised by medians with interquartile range (IQR) for non-normal distributions. Numbers and proportions are presented, and where data were missing, proportions are presented out of cases known. Statistical analyses were performed using STATA version 17 (Statacorp, TX, USA).

In this national observational study, the study sample size was governed by the disease incidence, thus no formal power calculation was carried out.

## RESULTS

Of the 1561 women admitted with confirmed SARS-CoV-2 infection between 15^th^ December 2021 and 14^th^ January 2022 (Figure 1), 449 (28.8%) were symptomatic and 1112 (71.2%) were asymptomatic.

The characteristics of included women stratified by admission group are shown in Table 1. The proportion of women aged 35 years or over was similar among symptomatic and asymptomatic women: 27.5% (n=115) and 26.3% (n=278), respectively. BMI of 30kg/m^2^ or more was reported for 29.7% (n=127) and 27.1% (n=289) among symptomatic and asymptomatic women, respectively. Black, Asian or other minority background was reported for 33.5% (n=147) and 38.4% (n=412). In the symptomatic group, 50.8% (n=224) had a gestational age at admission from 22 to 36 completed weeks; this proportion was 22.8% (n=227) in the asymptomatic group. Vaccination status was known for 1274 women (81.6%), 383 in the symptomatic group and 891 in the asymptomatic group.

**Table 1.** Sociodemographic characteristics and medical risk factors among pregnant women admitted with SARS-CoV-2, by admission group, UK, 15^th^ December 2021 to 14^th^ January 2022

| Characteristic | Symptomatic (n,%)<br>(n=449) | Asymptomatic (n,%)<br>(n=1112) |
| --- | --- | --- |
| Age (years): |  |  |
| <20 | 9 (2.2) | 31 (2.9) |
| 20-34 | 294 (70.3) | 750 (70.8) |
| ≥35 | 115 (27.5) | 278 (26.3) |
| Missing | 31 | 53 |
| Body Mass Index (BMI) (kg/m<sup>2</sup>): |  |  |
| Underweight (<18.5) | 10 (2.3) | 38 (3.6) |
| Normal (18.5 to <25) | 161 (37.6) | 417 (39.0) |
| Overweight (25 to <30) | 130 (30.4) | 324 (30.3) |
| Obese (>30) | 127 (29.7) | 289 (27.1) |
| Missing | 21 | 44 |
| Either woman or partner in paid work | 341 (76.0) | 841 (75.6) |
| Ethnic Group |  |  |
| White | 292 (66.5) | 661 (61.6) |
| Asian | 58 (13.2) | 179 (16.7) |
| Black | 53 (12.1) | 162 (15.1) |
| Chinese/Other | 12 (2.7) | 40 (3.7) |
| Mixed | 24 (5.5) | 31 (2.9) |
| Missing | 10 | 39 |
| Current smoking | 73 (16.9) | 190 (17.7) |
| Missing | 17 | 40 |
| Pre-existing medical conditions |  |  |
| Asthma | 40 (8.9) | 60 (5.4) |
| Hypertension | 10 (2.2) | 13 (1.2) |
| Cardiac disease | 4 (0.9) | 8 (0.7) |
| Diabetes | 9 (2.0) | 10 (0.9) |
| Multiparous | 271 (61.0) | 662 (60.9) |
| Missing | 5 | 25 |
| Multiple pregnancy | 5 (1.1) | 12 (1.1) |
| Gestation at admission (weeks) |  |  |
| <22 | 32 (7.3) | 67 (6.1) |
| 22-27<sup>+6</sup> | 40 (9.1) | 33 (3.0) |
| 28-33<sup>+6</sup> | 100 (22.7) | 81 (7.4) |
| 34-36<sup>+6</sup> | 84 (19.1) | 113 (10.3) |
| 37 or more | 185 (42.0) | 800 (73.1) |
| Missing | 8 | 18 |
| Vaccination status |  |  |
| Unvaccinated | 249 (65.1) | 625 (70.2) |
| 1 dose | 45 (11.8) | 83 (9.3) |
| 2 doses | 76 (19.8) | 154 (17.3) |
| 3 doses | 13 (3.4) | 29 (3.2) |
| Not known/<br>not documented | 66 | 221 |

### Respiratory support and medical treatment for symptomatic women

Two women (0.2%) in the asymptomatic group were admitted to ICU for indications unrelated to their SARS-CoV-2 infection. Overall, 86 (19.2%) of the symptomatic women had at least one indicator of moderate to severe infection (Table 2). However, the proportion of symptomatic women who received any specific pharmacological therapy was low (n=31, 6.9%); 0.4% (n=2) received antivirals, 1.6% (n=7) received tocilizumab, 5.8% (n=26) received corticosteroids for maternal indication, and 0.9% (n=4) received monoclonal antibodies. Four women were recruited to the RECOVERY trial. Among the 19 symptomatic women admitted to ICU, 10 (52.6%) received a specific pharmacological therapy, 1 woman (5.3%) received antivirals, 4 (21.0%) received tocilizumab, 9 (47.4%) received corticosteroids for a maternal indication and 2 (10.5%) received monoclonal antibodies.

**Table 2.** Respiratory support and medical treatment to symptomatic pregnant women admitted to hospital with SARS-CoV-2 infection, UK, 15^th^ December 2021 to 14^th^ January 2022

|  | Number of women<br>n (%) |
| --- | --- |
| Total number | 449 |
| Composite indicator of moderate to severe infection | 86 (19.2) |
| Oxygen saturation measured on admission (Yes) | 392 (87.3) |
| Oxygen saturation <95% | 18 (5.4) |
| Evidence of pneumonia on imaging | 51 (11.4) |
| Respiratory support required | 62 (14.3) |
| Non-invasive oxygen (nasal canulae, mask or non-rebreathe mask at <15l/min) | 40 (69.0) |
| High flow nasal oxygen (≥15l/min) or CPAP* | 8 (13.8) |
| Mechanic ventilation or ECMO<sup>†</sup> | 10 (17.2) |
| Level not known | 5 |
| Intensive Care Unit admission | 19 (4.2) |
| Maternal Death | 3 (0.7) |
| Pharmacological Management Total<sup>‡</sup> | 31 (6.9) |
| antivirals | 2 (0.4) |
| tocilizumab | 7 (1.6) |
| corticosteroids for maternal indication | 26 (5.8) |
| monoclonal antibodies | 4 (0.9) |
| recruited to RECOVERY-trial | 4 (0.9) |
| * CPAP: continuous positive airway pressure<br><sup>†</sup>ECMO: extracorporeal membrane oxygenation.<br><sup>‡</sup>Any of the listed medications given for medical management of SARS-CoV-2: antivirals, tocilizumab, maternal corticosteroids, monoclonal antibodies. |  |

**Table 3.** Outcomes among symptomatic pregnant women admitted to hospital with SARS-CoV-2 infection during the period when Omicron was the dominant variant by vaccination status, UK, 15^th^ December 2021 to 14^th^ January 2022

|  | Unvaccinated<br>n (%) | 1 dose<br>n (%) | 2 doses<br>n (%) | 3 doses<br>n (%) | Status<br>unknown n<br>(%) |
| --- | --- | --- | --- | --- | --- |
| Total number | 249 | 45 | (n=76) | (n=13) | (n=66) |
| Composite indicator of moderate to severe infection | 59 (23.7) | 10 (22.2) | 9 (11.8) | 0 (0.0) | 8 (12.1) |
| Intensive Care admission | 14 (5.6) | 3 (6.7) | 1 (1.3) | 0 (0.0) | 1 (1.5) |
| Maternal Death | 2 (0.8) | 1 (2.2) | 0 (0.0) | 0 (0.0) | 0 (0.0) |

### Vaccination status

The proportion of symptomatic women who had received none, one, two or three vaccine doses was 65.1% (n=249), 11.8% (n=45), 19.8% (n=76), and 3.4% (n=13), respectively (Table 1). A total of 78 (20.4%) symptomatic women whose vaccination status was known had a composite measure of moderate to severe infection. More than a fifth of unvaccinated women (59/249, 23.7%) and women who had received one dose (10/45, 22.2%) had moderate to severe infection, compared to one in ten (9/76, 11.8%) who had two doses and none (0/13, 0%) who had three doses. Forty of the women in the two-dose group (52.6%) were known to have received their second vaccine dose more than three months prior to admission; this included five of the nine women who had a composite indicator of moderate to severe infection and the woman who was admitted to ICU. Overall, none of the women (0/68) who had a composite indicator of moderate to severe infection and known vaccination status had completed the vaccination schedule advised to protect the general adult population against severe omicron infection.

### Pregnancy outcomes

One thousand, one hundred and seventy-two women had completed their pregnancies, 240 (53.5%) of the women in the symptomatic group and 932 (83.8%) in the asymptomatic group (Table 4). Almost a third of symptomatic women (n=144, 32.1%) were known to have been discharged still pregnant. The proportion of births at gestational weeks 22 to 36 was 17.8% (n=42) amongst symptomatic women versus 10.7% (n=96) in asymptomatic women. Birth expedited due to covid-19 was reported for 7.6% (n=18) in the symptomatic group; none of these women were known to have received three vaccine doses (Table 5).

**Table 4.** Pregnancy outcomes for women admitted with SARS-CoV-2 infection during the period when Omicron was the dominant variant, by cause of admission, UK, 15^th^ December 2021 to 14^th^ January 2022

| Pregnancy outcome | Symptomatic<br>n (%) | Asymptomatic<br>n (%) |
| --- | --- | --- |
| Total number | 449 | 1112 |
| Still pregnant, known to have been discharged | 144 (32.1) | 107 (9.6) |
| Still pregnant, not known to have been discharged | 65 (14.5) | 73 (6.6) |
| Pregnancy completed | 240 (53.5) | 932 (83.8) |
| Pregnancy Loss | 3 (0.7) | 27 (2.4) |
| Birth | 237 (52.8) | 905 (81.4) |
| Gestation at birth (weeks<sup>+days</sup>)* |  |  |
| 22<sup>+0</sup> – 27<sup>+6</sup> | 1 (0.4) | 6 (0.7) |
| 28<sup>+0</sup> – 33<sup>+6</sup> | 8 (3.4) | 22 (2.5) |
| 34<sup>+0</sup> – 36<sup>+6</sup> | 33 (14.0) | 68 (7.6) |
| 37<sup>+0</sup> or more | 194 (82.2) | 802 (89.3) |
| Missing | 1 | 7 |
| Birth expedited due to COVID-19* | 18 (7.6) | 0 (0.0) |
| Mode of birth* |  |  |
| Pre-labour Caesarean | 76 (32.1) | 213 (23.7) |
| Caesarean after labour onset | 45 (19.0) | 127 (14.1) |
| Operative vaginal | 24 (10.1) | 104 (11.6) |
| Unassisted vaginal | 92 (38.8) | 454 (50.6) |
| Missing | 0 | 7 |
| * Excluding pregnancy loss from denominator |  |  |

**Table 5.** Pregnancy outcomes for women admitted with SARS-CoV-2 infection during the period when Omicron was the dominant variant, by cause of admission and vaccination status, UK, 15^th^ December 2021 to 14^th^ January 2022.

|  | Symptomatic<br>(n=449) |  |  |  |  | Asymptomatic<br>(n=1,112) |  |  |  |  |
| --- | --- | --- | --- | --- | --- | --- | --- | --- | --- | --- |
| Vaccination status | Unvaccinated<br>n (%) | 1 dose only<br>n (%) | 2 doses<br>n (%) | 3 doses<br>n (%) | Status<br>unknown<br>n (%) | Unvaccinated<br>n (%) | 1 dose<br>n (%) | 2 doses<br>n (%) | 3 doses<br>n (%) | Status<br>unknown<br>n (%) |
| Total number | 249 | 45 | 76 | 13 | 66 | 625 | 83 | 154 | 29 | 221 |
| Still pregnant, known to have been discharged | 71 (28.5) | 16 (35.6) | 23 (30.3) | 6 (46.1) | 28 (42.4) | 47 (7.5) | 9 (10.8) | 15 (9.7) | 4 (13.8) | 32 (14.5) |
| Still pregnant, not known to have been discharged | 38 (15.3) | 3 (6.7) | 15 (19.7) | 0 (0.0) | 9 (13.6) | 50 (8.0) | 4 (4.8) | 8 (5.2) | 1 (3.4) | 10 (4.5) |
| Pregnancy completed | 140 (56.2) | 26 (57.8) | 38 (50.0) | 7 (53.9) | 29 (43.9) | 528 (84.5) | 70 (84.3) | 131 (85.1) | 24 (82.8) | 179 (81.0) |
| Pregnancy Loss | 2 (0.8) | 0 (0.0) | 1 (1.3) | 0 (0.0) | 0 (0.0) | 13 (2.1) | 2 (2.4) | 4 (2.6) | 2 (6.9) | 6 (2.7) |
| Birth | 138 (55.4) | 26 (57.8) | 37 (48.7) | 7 (53.9) | 29 (43.9) | 515 (82.4) | 68 (81.9) | 127 (82.5) | 22 (75.9) | 173 (78.3) |
| Gestation at birth<br>(weeks<sup>+days</sup>) * |  |  |  |  |  |  |  |  |  |  |
| 22<sup>+0</sup> – 27<sup>+6</sup> | 1 (0.7) | 0 (0.0) | 0 (0.0) | 0 (0.0) | 0 (0.0) | 4 (0.8) | 0 (0.0) | 0 (0.0) | 0 (0.0) | 2 (1.2) |
| 28<sup>+0</sup> – 33<sup>+6</sup> | 5 (3.6) | 1 (4.0) | 0 (0.0) | 0 (0.0) | 2 (6.9) | 7 (1.4) | 5 (7.3) | 7 (5.6) | 0 (0.0) | 3 (1.7) |
| 34<sup>+0</sup> – 36<sup>+6</sup> | 24 (17.4) | 0 (0.0) | 7 (18.9) | 0 (0.0) | 2 (6.9) | 41 (8.0) | 2 (2.9) | 8 (6.4) | 3 (13.6) | 14 (8.2) |
| 37<sup>+0</sup> or more | 108 (78.3) | 24 (96.0) | 30 (81.1) | 7 (100.0) | 25 (86.2) | 460 (89.7) | 61 (89.7) | 110 (88.0) | 19 (86.4) | 152 (88.9) |
| Missing | 0 | 1 | 0 | 0 | 0 | 3 | 0 | 2 | 0 | 2 |
| Birth expedited due to COVID-19* | 12 (8.7) | 1 (3.8) | 2 (5.4) | 0 (0.0) | 3 (10.3) | 0 (0.0) | 0 (0.0) | 0 (0.0) | 0 (0.0) | 0 (0.0) |
| Mode of birth* |  |  |  |  |  |  |  |  |  |  |
| Pre-labour Caesarean | 42 (30.4) | 12 (46.2) | 12 (32.4) | 1 (14.3) | 9 (31.0) | 110 (21.4) | 23 (34.9) | 34 (27.2) | 7 (31.8) | 39 (22.7) |
| Caesarean after labour onset | 28 (20.3) | 7 (26.9) | 4 (10.8) | 2 (28.6) | 4 (13.8) | 80 (15.6) | 7 (10.6) | 18 (14.4) | 0 (0.0) | 22 (12.8) |
| Operative vaginal | 11 (8.0) | 2 (7.7) | 6 (16.2) | 2 (28.6) | 3 (10.3) | 65 (12.7) | 2 (3.0) | 12 (9.6) | 3 (13.6) | 22 (12.8) |
| Unassisted vaginal | 57 (41.3) | 5 (19.2) | 15 (40.5) | 2 (28.6) | 13 (44.8) | 258 (50.3) | 34 (51.5) | 61 (48.8) | 12 (54.6) | 89 (51.7) |
| Missing | 0 | 0 | 0 | 0 | 0 | 2 | 2 | 2 | 0 | 1 |

\*Excluding pregnancy loss from denominator

Among 1159 infants, 10 stillbirths were reported; in the symptomatic and asymptomatic groups stillbirths occurred in 0.8% (n=2) and 0.9% (n=8) of total births, respectively (Table 6). Eight of the ten stillbirths occurred to women who were unvaccinated or had one vaccine dose, but the role of SARS-CoV-2 in the stillbirth needs to be assessed in formal audit. Admission to a neonatal unit was reported for 15.4% (n=37) of infants born to symptomatic women and 8.5% (n=78) of infants born to asymptomatic women.

**Table 6:** Perinatal outcomes for women admitted with SARS-CoV-2 infection during the period when omicron was the dominant variant and who have given birth (n=1159) by cause of admission and vaccination status, UK, 15^th^ December 2021 to 14^th^ January 2022.

|  | **Symptomatic**<br>**(n=242)** |  |  |  |  | **Asymptomatic**<br>**(n=917)** |  |  |  |  |
| --- | --- | --- | --- | --- | --- | --- | --- | --- | --- | --- |
| Vaccine status | Unvaccinated<br>n (%) | 1 dose<br>n (%) | 2 doses<br>n (%) | 3 doses<br>n (%) | Status unknown<br>n (%) | Unvaccinated<br>n (%) | 1 dose<br>n (%) | 2 doses<br>n (%) | 3 doses<br>n (%) | Status unknown<br>n (%) |
| Total number | 141 | 27 | 37 | 7 | 30 | 521 | 68 | 130 | 23 | 175 |
| Stillbirth | 1 (0.7) | 1 (3.7) | 0 (0.0) | 0 (0.0) | 0 (0.0) | 5 (1.0) | 1 (1.5) | 2 (1.5) | 0 (0.0) | 0 (0.0) |
| Admission to Neonatal Unit* | 14 (10.0) | 5 (19.2) | 8 (21.6) | 1 (14.3) | 9 (30.0) | 39 (7.6) | 5 (7.5) | 12 (9.5) | 3 (13.0) | 18 (10.3) |
| Neonatal Death | 0 (0.0) | 0 (0.0) | 0 (0.0) | 0(0.0) | 0(0.0) | 3 (0.6) | 0 (0.0) | 0 (0.0) | 0 (0.0) | 1 (0.6) |
| *One infant (asymptomatic mother) had missing data for admission to neonatal unit. |  |  |  |  |  |  |  |  |  |  |

## DISCUSSION

### Principal findings

This national prospective cohort study has identified that among pregnant women admitted with SARS-CoV-2 infection during the period when the Omicron VOC was dominant around one in four were symptomatic. One in seven of these symptomatic pregnant women needed respiratory support. One in four symptomatic pregnant women who had received no vaccine or a single dose had moderate to severe infection. One in eight symptomatic pregnant women who had received two doses had moderate to severe infection. No symptomatic pregnant women who had received three doses had moderate or severe infection, though the number of pregnant women admitted with symptomatic SARS-CoV-2 infection who had received three vaccine doses was very small. No women with moderate to severe respiratory disease, ICU admission or who died had received vaccines according to the recommended schedule for the general adult population for the Omicron variant.

### Strengths and weaknesses of the study

To our knowledge, this is the first national prospective cohort study to describe pregnancy and perinatal outcomes during the period when the Omicron SARS-CoV-2 variant was dominant. A key strength of these data is the existing mechanism for national case identification of all women admitted to hospital across the UK, and therefore the low risk of selection bias. In the UK, universal SARS-CoV-2 testing for all obstetric admissions was implemented from May 2020. Asymptomatic pregnant women in whom SARS-CoV-2 infection is detected by screening on admission, are most commonly admitted to give birth.^17^ We therefore categorised the included women by cause of admission or symptoms to avoid misclassification bias and increased adverse outcomes being incorrectly attributed to SARS-CoV-2.^18^

Some of the pregnant women who had received two vaccine doses or fewer may also have delayed the second dose due to covid-19 infection; information about previous infection was not available in the current study. These women could potentially be misclassified into a category with lower expected protection while having reduced risk due to post-infection immunity, and this could result in overestimation of the protective effect of different vaccine doses. As with previous analyses,^5^ variant sequencing data were not available for individual women, and a proxy time period was used instead which may be considered a limitation. Additionally, more women in the symptomatic group have not completed their pregnancies, compared to the asymptomatic group, which is likely to affect the observed rates of key neonatal outcomes.

### Interpretation and comparison with related studies

The proportion of symptomatic women with moderate to severe infection was lower overall than in the wildtype, Alpha and Delta variant periods in the UK^5 13^. However, a greater proportion of symptomatic pregnant women had received one or more vaccine doses than in previous variant periods and this needs to be taken into account when comparing outcomes across variant periods, recognising that prior vaccination is likely to confer some degree of protection from both severe illness and symptomatic infection. When solely unvaccinated pregnant women admitted with symptomatic infection are considered, maternal outcomes are very similar to those observed during the initial wildtype infection period^5^. Among those in need of respiratory support, irrespective of vaccination status, the use of mechanic ventilation or ECMO was 16.1% (10/62) and thus lower than previous periods (23.5% in Alpha and 21.4% in Delta periods)^5^.

Covid-19-specific pharmacological therapies, which are now standard care for non-pregnant patients, were used infrequently, even for women that needed respiratory support. The proportion that received any pharmacological treatment for covid-19 (one or more of an antiviral, tocilizumab, maternal corticosteroids and monoclonal antibodies) was lower than in the Alpha and Delta periods, 6.9% vs 14.9% and 13.6% respectively. While this may partly reflect a lower severity of illness, it is concerning that only around half of pregnant women admitted to ICU due to covid-19 received any covid-19 specific pharmacological treatment. The RCOG recommended in June 2020 that corticosteroid therapy should be considered for all women who were clinically deteriorating due to covid-19.^16^ Maternal corticosteroid treatment was reported for 5.8% of symptomatic women during the Omicron period, compared to 12.7% and 12.0% during the Alpha and Delta periods, respectively. In the current study 47% of women admitted to intensive care received corticosteroids. Understanding this persisting low use of evidence-based therapies amongst severely ill pregnant and postpartum women is an increasingly urgent priority.

Few pregnant women who had received two or more doses of vaccine were admitted with symptomatic SARS-CoV-2, and none of the women with a composite indicator for moderate or severe infection had received three vaccine doses according to current recommendations to protect non-pregnant adults against severe omicron infection. Vaccination for all pregnant women regardless of risk group in the UK was recommended from 16^th^ April 2021, and all adults were eligible to receive vaccination from mid-June 2021.^19^ Vaccine coverage surveillance among women who gave birth in England up to October 2021 reported that 29.4% of the women had received two doses of vaccine and 58.2% were unvaccinated^20^. Similarly, vaccine coverage has been low in Scotland where 32.2% of women who gave birth in October 2021 had received two doses of vaccine during pregnancy compared to 77.4% of women of reproductive age (18-44 years), and 98.1% of women admitted to the ICU were unvaccinated.^6^ Almost 68% of the women with information about vaccination status included in the current study were unvaccinated.

In the general adult population, effectiveness after the second dose declines from 60-75% three weeks after vaccination to 20% at 15 weeks and 10% after 25 weeks,^20^ and three doses have been shown to give better protection against severe disease with the Omicron VOC in adults.^21^ The interval between the last dose and the admission was 3 months or more among half (53%) of the women who had received two doses of vaccine. The number of pregnant women who had received a third booster dose was low in our study, but no severe cases in this group indicates the importance of the third dose to protect pregnant women from both hospital admission with symptomatic covid-19 and need for respiratory support.

Disproportionate admissions due to covid-19 among pregnant women with ethnic minority backgrounds were less prominent in the current study than previously described during the wildtype period.^13^ National guidance has emphasised the importance of addressing this inequality and advised active health seeking in these groups.^16^ The observation time in the current study is short and the findings cannot yet reliably indicate if the smaller differences can be attributed to better communication, prevention, health care seeking strategies or previous infection. Preliminary surveillance results indicated that the Omicron VOC has a secondary attack rate of 10-13% and therefore factors that increase transmission, such as multi-occupancy housing and public-facing occupations, are important also for this variant.^22-24^ Since socioeconomic deprivation is also a known independent risk factor for adverse pregnancy outcome, this could be a source of residual confounding in this study.

Neonatal outcomes were purposely not compared between omicron and other periods as a high proportion of pregnancies were continuing at the time of analysis. However, the available data suggest that the risk of stillbirth during this period may be lower than observed during the delta period^5^. Further follow-up is required to clarify the effect of infection during the Omicron dominant period on perinatal outcomes such as stillbirth.

### Implications for clinicians and policymakers

The findings of this study indicate that the risk of severe respiratory failure in unvaccinated pregnant women with Omicron VOC is similar to that observed in the UK during the initial wildtype variant wave of the pandemic.^13^ While severe outcomes were less frequent in the current period compared to the previous Alpha and Delta variant dominant periods, it is important to keep in mind that the risk of hospital admission due to covid-19 was higher in the UK than in other European countries during the initial months of the pandemic,^25 26^ possibly associated with early implementation of public health measures to limit viral transmission. If public health interventions could to some extent protect pregnant women during the first wave, individual protection through vaccination is now available. Our results indicate that most current cases of respiratory failure among pregnant women are preventable, yet vaccine uptake among pregnant women remains low compared to the general female population in fertile age. Continued, strong efforts to improve uptake of the vaccine during pregnancy are still needed. This is of even greater importance as infection continues to rapidly rise in both high and low-resourced settings.^27^

## Data Availability

Data Sharing
Data cannot be shared publicly because of confidentiality issues and potential identifiability of sensitive data as identified within the Research Ethics Committee application/approval. Requests to access the data can be made by contacting the National Perinatal Epidemiology Unit data access committee via.

## Data Sharing

Data cannot be shared publicly because of confidentiality issues and potential identifiability of sensitive data as identified within the Research Ethics Committee application/approval. Requests to access the data can be made by contacting the National Perinatal Epidemiology Unit data access committee via.

## Acknowledgements

The authors would like to acknowledge the assistance of UKOSS reporting clinicians, the NIHR Reproductive Health & Childbirth National Research Champions, and the UKOSS Steering Committee without whose support this research would not have been possible.

## Contributorship statement

All authors contributed to conceptualisation, the writing and editing of this study, had final approval of the version to be published and agree to be accountable for all aspects of the work. KB, EM, NS, CG, PO, MQ, PB, JK and MK contributed to funding acquisition, supervision, and methodology. HE, RR, NV, KB and MK contributed to data curation and formal analysis, and had access to verify the underlying data. MK, as guarantor, accepts full responsibility for the work and affirms that the manuscript is an honest, accurate and transparent account of the study being reported; that no important aspects of the study have been omitted; and that any discrepancies from the study as originally planned have been explained. The corresponding author attests that all listed authors meet authorship criteria and that no others meeting the criteria have been omitted.

The Corresponding Author has the right to grant on behalf of all authors and does grant on behalf of all authors, a worldwide licence to the Publishers and its licensees in perpetuity, in all forms, formats and media (whether known now or created in the future), to i) publish, reproduce, distribute, display and store the Contribution, ii) translate the Contribution into other languages, create adaptations, reprints, include within collections and create summaries, extracts and/or, abstracts of the Contribution, iii) create any other derivative work(s) based on the Contribution, iv) to exploit all subsidiary rights in the Contribution, v) the inclusion of electronic links from the Contribution to third party material where-ever it may be located; and, vi) licence any third party to do any or all of the above.

